# rs641738C>T near *MBOAT7* is positively associated with liver fat, ALT, and histological severity of NAFLD: a meta-analysis

**DOI:** 10.1101/19013623

**Authors:** Kevin Teo, Kushala W. M. Abeysekera, Leon Adams, Elmar Aigner, Jesus M. Banales, Rajarshi Banerjee, Priyadarshi Basu, Thomas Berg, Pallav Bhatnagar, Stephan Buch, Ali Canbay, Sonia Caprio, Ankita Chatterjee, Yii-Der Ida Chen, Abhijit Chowdhury, Christian Datz, Dana de Gracia Hahn, Johanna K. DiStefano, Jiawen Dong, Amedine Duret, EU-PNAFLD Investigators, Connor Emdin, Madison Fairey, Glenn S Gerhard, GOLD Consortium, Xiuqing Guo, Jochen Hampe, Matthew Hickman, Lena Heintz, Christian Hudert, Harriet Hunter, Matt Kelly, Julia Kozlitina, Marcin Krawczyk, Frank Lammert, Claudia Langenberg, Joel Lavine, Lin Li, Hong Kai Lim, Rohit Loomba, Panu K. Luukkonen, Phillip E. Melton, Trevor A. Mori, Nicholette D. Palmer, Constantinos A. Parisinos, Sreekumar G Pillai, Faiza Qayyum, Matthias C. Reichert, Stefano Romeo, Jerome I. Rotter, Yu Ri Im, Nicola Santoro, Clemens Schafmayer, Elizabeth K. Speliotes, Stefan Stender, Felix Stickel, Christopher D. Still, Pavel Strnad, Kent D. Taylor, Anne Tybjærg-Hansen, Giuseppina Rosaria Umano, Mrudula Utukuri, Luca Valenti, Lynne E. Wagenknecht, Nicholas J. Wareham, Richard M. Watanabe, Julia Wattacheril, Hanieh Yaghootkar, Hannele Yki-Järvinen, Kendra A. Young, Jake P. Mann

## Abstract

**Background & Aims:** A common genetic variant near *MBOAT7* (rs641738C>T) has been previously associated with hepatic fat and advanced histology in non-alcoholic fatty liver disease (NAFLD), however, these findings have not been consistently replicated in the literature. We aimed to establish whether rs641738C>T is a risk factor across the spectrum of NAFLD and characterize its role in the regulation of related metabolic phenotypes through meta-analysis.

**Methods:** We performed meta-analysis of studies with data on the association between rs641738C>T genotype and: liver fat, NAFLD histology, and serum ALT, lipids, or insulin. These included directly genotyped studies and population-level data from genome-wide association studies (GWAS). We performed random effects meta-analysis using recessive, additive, and dominant genetic models.

**Results:** Data from 1,047,265 participants (8,303 with liver biopsies) across 42 studies was included in the meta-analysis. rs641738C>T was associated with higher liver fat on CT/MRI (+0.03 standard deviations [95% CI: 0.02 - 0.05]) and diagnosis of NAFLD (OR 1.22 [95% CI 1.08 - 1.39]) in Caucasian adults. The variant was also positively associated with presence of severe steatosis, NASH, and advanced fibrosis (OR: 1.32 [95% CI: 1.06 - 1.63]) in Caucasian adults using a recessive model of inheritance (CC+CT vs. TT). Meta-analysis of data from previous GWAS found the variant to be associated with higher ALT (P_z_=0.002) and lower serum triglycerides (P_z_=1.5×10^−4^). rs641738C>T was not associated with fasting insulin and no effect was observed in children with NAFLD.

**Conclusion:** Our study validates rs641738C>T near *MBOAT7* as a risk factor for the presence and severity of NAFLD in individuals of European descent.

## INTRODUCTION

Since the first genome-wide association study (GWAS) of liver fat[1], more than 20 genetic single nucleotide variants (SNVs) have been associated with non-alcoholic fatty liver disease (NAFLD)[2,3]. These studies have deepened our understanding of the condition, its heritability, and its relationship with cardio-metabolic disease.

Rs641738C>T near *MBOAT7* (membrane bound O-acyltransferase domain containing 7) was initially identified as a genome-wide significant risk variant for alcohol-related cirrhosis (odds ratio=1.35, p=1.03 × 10−9)) [4]. It has since been implicated in the pathogenesis of NAFLD[5], hepatocellular carcinoma[6], as well as in fibrosis development in chronic hepatitis B/C[7–9], and primary sclerosing cholangitis[10]. However, unlike variants in *PNPLA3, TM6SF2*, and *HSD17B13*, it has not been identified at genome-wide significance for liver fat or ALT[1,11,12].

Rs641738 is located a few hundred base pairs downstream of the 3’ untranslated region of *MBOAT7*, which belongs to a family of genes that code for specific acyl donors and acceptors[13]. *MBOAT7* encodes lysophosphatidylinositol acyltransferase 1 (LPIAT1), which contributes to the regulation of free arachidonic acid in cells[14,15]. Rs641738C>T is associated with lower hepatic expression of *MBOAT7* at both the mRNA[16] and protein level[5]. Given its role in inflammatory lipid pathways, most mechanistic work relating to rs641738 has focused on *MBOAT7[17]*.

In NAFLD, the rs641738C>T variant was first demonstrated to be associated with increased hepatic fat content and severity of fibrosis in individuals of European descent[5]. Proton magnetic resonance spectroscopy data from 2736 individuals showed a modest increase in hepatic fat in those with TT-genotype (4.1%) compared to those with CT-(3.6%) or CC-genotype (3.5%, p=.005). Follow-up studies of European subjects corroborated the initial findings, and suggested a role in development of hepatocellular carcinoma[18,19]. However, these results were not replicated in adults of other ancestries[5,20–22] or in children[23].

In addition, biallelic loss of function mutations in *MBOAT7* cause autosomal recessive mental retardation 57 (OMIM #617188) and no liver phenotype has been reported in these patients to date[15,24,25]. However, rare likely pathogenic (coding) variants in *MBOAT7* are associated with HCC in NAFLD[26].

In summary, the association between rs641738C>T and hepatic fat content, as well as its effects on severity of NAFLD, remain unclear. Moreover, the broader metabolic effects of this SNV, including its association with markers of insulin resistance and dyslipidaemia have not been assessed. Understanding the broader metabolic effects of rs641738C>T is important if it were to be investigated as a drug target in NAFLD.

Here, we conducted a large meta-analysis to determine if rs641738C>T influences the development or stage of NAFLD and related traits.

## METHODS

### Data sources and study selection

Two data sources were included in the meta-analysis: (i) studies which looked at the effect of the variant on traits of interest by genotyping the variant; and (ii) look-up from GWAS of traits of interest.

Published studies were sourced through: Medline, Embase, HuGe Navigator, Web of Science, bioRxiv, and medRxiv. The search terms used were: “(*MBOAT7* or membrane-bound-o-acyltransferase) or (rs641738 or rs626283) or (*TMC4*)”. In addition, HuGe Navigator Phenopedia was searched using terms related to liver disease (Supplementary Methods). There were no restrictions on date or language, and the study selection included all original studies including AASLD Liver Meeting and EASL meeting abstracts. The search was completed on 4^th^ March 2020. Reference lists of publications were also reviewed.

A separate search was conducted for all potentially relevant GWAS through: GWAS Catalogue[27], Phenoscanner[28], Type 2 diabetes knowledge portal[29], and Cardiovascular disease knowledge portal[30] (Supplementary Methods).

After removal of duplicates, titles and abstracts were screened for eligibility independently by two authors (investigators), with inclusion/exclusion criteria applied to potentially eligible full texts.

HuGENet guidelines[31] were followed throughout and MOOSE reporting guidelines[32] were used. This study was prospectively registered on PROSPERO Database of Systematic Reviews (CRD42018105507) Available from: http://www.crd.york.ac.uk/PROSPERO/display_record.php?ID=CRD42018105507

### Inclusion and exclusion criteria

Studies were included if genotyping of rs641738C>T (or rs626283G>C [R^2^>0.98 in European and American populations[33]] / rs2576452C>T [R^2^=0.92 in Guzman *et al*. [34]], which are in strong linkage disequilibrium with rs641738C>T) was conducted and data on one of the outcomes of interest were reported. Narrative review articles, *in vitro* studies, and investigations involving animals, fish, and invertebrates were excluded. Studies which investigated liver disease of other etiologies were also excluded. There was no restriction on ethnicity or ancestry. Types of studies eligible for inclusion were: case-control, cohort, genome-wide association studies, systematic reviews, and meta-analyses. Pre-print and abstract publications were eligible for inclusion provided that sufficiently detailed information could be obtained for assessment of inclusion/exclusion criteria, through availability of individual participant-level data, where possible. Several studies reported on the same cohort (or patient sample) in more than one article. In these instances, data only from the larger of the overlapping cohorts were included in analyses. A full list of overlapping cohorts and articles is in Supplementary Table 1.

### Outcomes of interest

Given the strong evidence base for fibrosis stage as a prognostic marker in NAFLD[35], the primary outcome for the meta-analysis was the association of rs641738C>T with presence of advanced fibrosis (F3-4 versus F0-2) in individuals with NAFLD.

Dichotomous secondary outcomes were: radiological diagnosis of NAFLD (or hepatic steatosis); presence of severe steatosis on liver biopsy (S3 versus S1-2); presence of non-alcoholic steatohepatitis (NASH) on liver biopsy (NASH versus non-alcoholic fatty liver (NAFL)); presence of any fibrosis (F1-4 versus F0); presence of hepatocellular carcinoma (HCC) in patients with NAFLD (NAFLD-HCC versus NAFLD).

Continuous secondary outcomes were: quantitative, radiological hepatic fat content; total cholesterol; high-density lipoprotein cholesterol (HDL); low-density lipoprotein cholesterol (LDL); triglycerides (TG).

### Data collection

Details on the recruitment of controls and cases were obtained from each study and, where necessary, clarified by discussion with the study’s authors. In particular, it was noted when cases and controls were not recruited from the same population or clinics.

Hepatic steatosis or NAFLD (as diagnosis) was evaluated as a dichotomous variable where radiological assessment (liver ultrasound, controlled attenuation parameter [CAP, with cut-off >240m/s], CT, MRI) were used. Hepatic fat content was collected as a continuous variable from CT, MRS, MRI, PDFF. Non-invasive assessment of hepatic fat content was also assessed using semi-quantitative scoring in the Fenland cohort, as previously described[36], and using CAP.

Individual participant-level histology data were extracted according to the NASH Clinical Research Network scoring system[37] and, where not otherwise diagnosed by a pathologist’s assessment, NASH was defined using the Fatty Liver Inhibition of Progression (FLIP) algorithm[38]. The above data were collected for each genotype separately (CC, CT, and TT).

Participant demographics and characteristics meta-data were collected from each study, including: sex, age, ethnicity, presence of type 2 diabetes, body mass index (BMI). Where possible, individual patient-level data was obtained. The authors of 58 studies were contacted for additional data or clarification, of whom 48 replied. Data from 15 potentially relevant studies could not be included, which are listed in the Supplementary Methods.

### Cohorts with genome-wide data

Relevant GWASs were identified as described above. Data were included where densely imputed genotyping results were available for rs641738C>T or rs626283C>G in all with >0.98 call rate. These cohorts have been described elsewhere and detailed description of the quality control processes for genome-wide data is available in their original descriptions (Table 1), but in brief, single nucleotide polymorphisms (SNP) with Hardy-Weinberg equilibrium p-values <1×10^−6^ were excluded prior to imputation. Association analysis was performed for variants with mean allele frequencies >0.01 and with minimum imputation quality of >0.3.

**Table 1.** Characteristics of studies included in the meta-analysis. Further characteristics available in Supplementary Tables 2-3. ALSPAC, Avon Longitudinal Study of Parents and Children; CAP, controlled attenuation parameter; CT, computerized tomography; DHS, Dallas Heart Study; GWAS, genome-wide association study; LB, liver biopsy; LBC, Liver Biopsy Cohort; MRI, magnetic resonance imaging; MRS, magnetic resonance spectroscopy; NA, not applicable; US, ultrasound.

| Study;<br>Age group | Study design<br>and sample size<br>(N) | Liver<br>biopsy<br>(N) | Features and participant<br>characteristics | Outcomes |
| --- | --- | --- | --- | --- |
| Abeysekera<br>2019[46];<br>Adult | Population-based<br>cohort<br>N=2,919 | NA | GWAS data from the ALSPAC birth cohort study with steatosis diagnosed using CAP >240kPa | NAFLD diagnosis, liver fat (CAP) |
| Adams<br>2013[47];<br>Paediatric | Population-based<br>cohort<br>N=948 | NA | GWAS of adolescents with hepatic steatosis measured by US | NAFLD diagnosis, biochemistry |
| Caussy<br>2019[48];<br>Adult | Family, twin, &<br>sibling-paired<br>study<br>N=104 | NA | Community-based family case-control study with NAFLD diagnosed by MRI-PDFF | NAFLD diagnosis, liver fat, biochemistry |
| Chambers<br>2011[49];<br>Adult | Population-based<br>cohorts<br>N=61,089 | NA | GWAS of adults from 21 population-based cohorts for concentration of serum liver enzymes | ALT (GWAS) |
| Chatterjee<br>2018[50];<br>Adult | Hospital-based<br>case-control<br>N=354 | 132 | Exome-wide association study of adults with NAFLD diagnosed by LB or MRI | NAFLD diagnosis, liver fat, histology |
| Chen 2020[51];<br>Adult | Hospital-based<br>cohort<br>N=19,598 | NA | GWAS of adults undergoing elective surgery with linkage to electronic health records | Biochemistry (GWAS) |
| Di Costanzo<br>2018[52];<br>Adult | Hospital-based<br>case-control<br>N=445 | NA | Sample from adult obesity clinic with NAFLD diagnosed using US | NAFLD diagnosis, biochemistry |
| Di Costanzo<br>2019[53];<br>Paediatric | Hospital-based<br>case-control<br>N=230 | NA | Sample from paediatric obesity clinic with NAFLD diagnosed using MRI hepatic fat fraction | NAFLD diagnosis, liver fat, biochemistry |
| Di Sessa<br>2018[54];<br>Paediatric | Hospital-based<br>case-control<br>N=1,002 | NA | Sample from paediatric obesity clinic with NAFLD diagnosed using US | NAFLD diagnosis, biochemistry |
| DiStefano<br>2015[55];<br>Adult | Hospital-based<br>case-control<br>N=1,868 | 1868 | GWAS of adults undergoing bariatric surgery with NAFLD diagnosed by LB | NAFLD diagnosis, histology |
| Donati 2017[6];<br>Adult | Hospital-based<br>case-control<br>N=1073 (Italian) | 1123 | Cohorts from adult liver/obesity clinics with NAFLD diagnosed using US and/or LB | NAFLD diagnosis, histology, HCC |
|  | N=358 (UK) |  |  |  |
| Dongiovanni 2018 (DHS)[56]; Adult | Population-based cohort<br>N=2,675 | NA | GWAS of adults from population-based cohort with NAFLD diagnosed using MRS | NAFLD diagnosis, liver fat, biochemistry |
| Dongiovanni 2018 (LBC)[56]; Adult | Hospital-based case-control<br>N=1,515 (LBC) | 1515 | Samples from adult liver/obesity clinics and bariatric surgery with NAFLD diagnosed using LB | NAFLD diagnosis, histology, biochemistry |
| Gurdasani 2019[57]; Adult | Population-based cohorts<br>N=13,116 | NA | GWAS of adults from 4 population-based cohort studies, including concentration of serum liver enzymes and biochemistry | Biochemistry (GWAS) |
| Guzman 2018[34]; Adult | Clinical trial-based case-control<br>N=148 | NA | Sample of adults with type 2 diabetes who had taken part in one of two interventional diabetes trials, with NAFLD diagnosed on MRI | Liver fat |
| Hudert 2019[23]; Paediatric | Hospital-based case-control<br>N=270 | 70 | Cases from paediatric liver clinic with diagnosis of NAFLD using LB | NAFLD diagnosis, liver fat, histology |
| Kanai 2018[58]; Adult | Population-based cohort<br>N=134,182 | NA | GWAS of adults from a population cohort study with biobank, including concentration of serum liver enzymes and biochemistry | Biochemistry (GWAS) |
| Karajamaki 2019[59]; Adult | Population-based case-control<br>N=935 | NA | Population-level case-control study with recruitment of cases with hypertension; NAFLD diagnosed using US | NAFLD diagnosis, biochemistry |
| Kawaguchi 2018[60]; Adult | Mixed hospital- and population-based case-control<br>N=8,571 | 936 | Sample of adults from liver clinics with NAFLD diagnosed by LB | NAFLD diagnosis, histology, HCC, biochemistry |
| Koo 2020[61]; Adult | Hospital-based case-control<br>N=461 | 461 | Sample of adults from hospital clinics with NAFLD diagnosed by LB | NAFLD diagnosis, histology, biochemistry |
| Krawczyk 2016[62]; Adult | Hospital-based case-control<br>N=84 | 84 | Samples from adults undergoing bariatric surgery with NAFLD diagnosed using LB | Histology, biochemistry |
| Krawczyk 2018[63]; Adult | Hospital-based case-control<br>N=515 | 295 | Sample of adults from liver clinics with NAFLD diagnosed by US or LB | Histology, liver fat (CAP), biochemistry |
| Lin 2018[64]; Paediatric | Population-based cohort<br>N=819 | NA | Cohort of children recruited from schools with diagnosis of NAFLD using US | NAFLD diagnosis, biochemistry |
| Luukkonen 2020[65];<br>Adult | Hospital-based case-control<br>N=793 | 236 | Sample of adults undergoing bariatric surgery with NAFLD diagnosed by LB or MRS | NAFLD diagnosis, liver fat, histology, biochemistry |
| Mann 2018a[66];<br>Adult | Population-based cohort<br>N=10,934 | NA | Cohort study of adults from population with NAFLD diagnosed by US | NAFLD diagnosis, liver fat (US), biochemistry |
| Mann 2018b[67];<br>Paediatric | Hospital-based case-control<br>N=379 | 306 | Sample from paediatric obesity and liver clinics with diagnosis of NAFLD using LB or US | NAFLD diagnosis, histology, biochemistry |
| Middelberg 2011[68];<br>Adult & paediatric | Population-based cohort<br>N=11,693 | NA | Combination of two cohort studies: adolescent twins and siblings; and adult twins, both population-based, with GWAS including concentration of serum liver enzymes and biochemistry | Biochemistry (GWAS) |
| Moon 2019[69];<br>Adult | Population-based cohort<br>N=6,949 | NA | GWAS of adults from a population cohort study with biobank, including concentration of serum liver enzymes and biochemistry | Biochemistry (GWAS) |
| Prins 2017[70];<br>Adult | Population-based cohort<br>N=9,731 | NA | GWAS of adults from a population cohort study with biobank, including concentration of serum liver enzymes and biochemistry | Biochemistry (GWAS) |
| Reichert 2019[71];<br>Adult | Hospital-based case-control<br>N= 54 | NA | Sample of adults from liver clinics with NAFLD cirrhosis diagnosed by LB, or US/MRI/CT | HCC |
| Seidelin 2020[72];<br>Adult | Population-based cohort<br>N=7511 | NA | Hepatic steatosis measured by CT, part of the Copenhagen General Population Study | Liver fat |
| Sookoian 2018[20];<br>Adult | Hospital-based case-control<br>N=634 | 372 | Samples from adult liver clinics and bariatric surgery patients with NAFLD diagnosed by LB or US | NAFLD diagnosis, histology |
| Speliotes 2011[1];<br>Adult | Population-based cohorts<br>N=4,244 | NA | GWAS of adults from 4 population-based cohorts for hepatic fat content as determined by CT | Liver fat |
| Strnad 2018[73];<br>Adult | Hospital-based case-control<br>N=672 | 672 | Sample of adults from liver clinics with NAFLD diagnosed by LB | Histology |
| Tabassum 2019[74];<br>Adult | Population-based cohorts<br>N=2,181 | NA | GWAS of adults from 2 population-based cohorts with data on serum triglycerides | Biochemistry (GWAS) |
| UKBB 2019[75];<br>Adult | Population-based<br>N=14,440 (liver fat); N=77,464 (coded fibrosis); N= 353,596 | NA | GWAS of hepatic steatosis measured by MRI from the UK BioBank; adults with coded diagnosis of NAFLD and cirrhosis; and concentration of serum | Liver fat, biochemistry (GWAS), fibrosis (coding data) |
|  | (biochemistry) |  | biochemistry available from GWAS |  |
| Umano 2018[22];<br>Paediatric | Hospital-based case-control<br>N=878 | NA | Sample from paediatric obesity clinic with NAFLD diagnosed using MRI hepatic fat fraction | Liver fat, biochemistry |
| Verma 2017[76];<br>Adult | Hospital-based cohort<br>N=31,466 | NA | GWAS of adult hospital patients with linkage to electronic health records | Biochemistry (GWAS) |
| Viitasalo 2016[77];<br>Paediatric | Population-based cohort<br>N=467 | NA | Population cohort of children recruited from schools with measurement of ALT | Biochemistry |
| Wattacheril 2017[78];<br>Paediatric | Cases-only<br>N=208 | 208 | GWAS of Hispanic boys with NAFLD diagnosed by LB | Histology |
| Willer 2013[79];<br>Adult | Population-based cohorts<br>N=347,532 | NA | GWAS of adults from 45 population-based cohorts with data on serum lipids | Biochemistry (GWAS) |
| Young 2019[80];<br>Adult | Population-based cohorts<br>N=3,555 | NA | GWAS of adults from 5 population-based cohorts with data on ALT | Biochemistry (GWAS) |
| Zusi 2019[81];<br>Paediatric | Hospital-based case-control<br>N=510 | NA | Sample from paediatric obesity clinic with diagnosis of NAFLD using US | NAFLD diagnosis, biochemistry |

Liver fat data from the UK BioBank was extracted under Application ID 9914 (‘Determining the Outcomes of People with Liver Disease’). Additional details regarding the Avon Longitudinal Study of Parents and Children (ALSPAC) [39–41] and data extracted from the UK BioBank cohort is found in the Supplementary Methods.

### Patient and Public Involvement

Patients and public were not directly involved in the planning of this meta-analysis; however, many of the original studies included were co-produced with patient and public involvement.

### Study quality assessment

Two reviewers independently assessed risk of bias in each study by applying the Cochrane Risk of Bias in Cohort Studies tool[42].

### Statistical Analysis

Genotype frequencies for each study were assessed for Hardy-Weinberg equilibrium using chi-squared test.

Due to the unclear effect of this variant on liver disease and previous studies using multiple different models of inheritance, genetic association analyses were performed using additive, dominant, and recessive models for each outcome.

For dichotomous outcomes, the effect statistic was calculated as an odds ratio between groups.

For analysis of diagnosis of liver fat, a sensitivity analysis was performed by excluding studies where there was a risk of confounding due to differences between cases and controls.

For analysis of effect on liver fat (continuous quantitative liver fat data from CT, MRI, MRS, or PDFF and semi-quantitative data using ultrasound or CAP), data were inverse normalized and an additive genetic model (coding the number of T alleles as 0, 1, and 2) was used with linear regression, adjusted for age, sex, and (where available) principal components of genetic ancestry. For other continuous variables where raw data was available (i.e. triglycerides, HDL, LDL, ALT, and total cholesterol), effect summary was calculated as a mean difference between CC and TT groups.

For meta-analysis of GWAS summary statistics, pooled effect summaries were calculated where traits had been logarithmically transformed and an additive model had been used in analyses. Beta regression coefficients were pooled using inverse-variance pooling with Fisher’s z-transformation. Meta-analysis was performed using random effects throughout using DerSimonian-Laird method for estimation of tau^2^.

Summary statistics were reported with 95% confidence intervals (CI). Data from paediatric and adult studies were analyzed separately. Sub-analysis was performed using only studies conducted in Caucasian populations (self-reported white, Non-Finnish-, or Finnish-European ethnicity) where data were available from at least four studies. This sub-analysis was selected due to initial identification of this variant in Caucasian individuals and further sub-analysis by ethnicity may be affected by differences in linkage disequilibrium between genetic ancestries. For studies including cohorts of multiple ethnicities, where available, data were analysed separately for each ethnicity. Heterogeneity between groups was described using the Q statistic, tau^2^, and I^2^. Where there was substantial heterogeneity (I^2^>50%, P_Q_<0.05) that remained after sub-group analysis by non-Caucasian population, univariable random-effects meta-regression was performed using: proportion of female participants, mean age, proportion with type 2 diabetes, and mean BMI. In addition, for the analysis of hepatic fat as a quantitative trait, meta-regression was performed for the imaging modality used (e.g. MRI-PDFF, MRS). For dichotomous outcomes: p-value<0.017 (i.e. p<0.05/3) was considered statistically significant due to testing outcomes against three genetic association models. For other outcomes, where only a single genetic model was used, p-value<0.05 was considered significant.

Bias was assessed using Egger’s test and funnel plots where more than 10 studies were included. Where Egger’s test suggested bias (p<0.05), a funnel plot was generated and missing studies were imputed using Duval & Tweedie’s trim-and-fill procedure[43].

Analysis was performed using STATAv14 for Windows (StataCorp. 2015. Stata Statistical Software: Release 14. College Station, TX: StataCorp LP), GraphPad Prism (v8.0 for Mac, GraphPad Software, La Jolla California, USA), and R 3.6.1[44,45]. Script used in analyses in R is available in the Supplementary Data.

## RESULTS

Database search identified 1120 articles (Supplementary Fig. 1), of which 43 articles were included: 42 primary studies (Table 1 & Supplementary Tables 2-3) and one meta-analysis (Supplementary Table 4).

In total, 1,047,265 individuals (5,711 children) were included in the meta-analysis. Most studies were in adults (32/42, 76%) and in individuals from predominantly Caucasian populations (26/42, 62%). Of the 42 included studies, 14 studies (8,303 participants, hereof 584 children) reported data on liver histology.

Studies were generally of high quality, though in four studies[23,52,60,61] (three in adults and one in children) the control group was recruited from a different population or sample to the cases (Supplementary Table 2).

One previous meta-analysis was included[82], which used data from 5 case-control studies to assess the effect of rs641738C>T on diagnosis of NAFLD. The meta-analysis included 2560 cases and 8738 controls and found no evidence of an association between this variant and diagnosis of NAFLD (Supplementary Table 4).

### Liver fat, NAFLD, and severe steatosis in adults

Nine studies (29,916 participants) reported data on hepatic fat as a continuous variable assayed by CT or MRI. On meta-analysis, rs641738C>T was associated with higher liver fat in studies in Caucasian populations, with a per T-allele change of β 0.034 (95% CI 0.018, 0.051), P_z_=5.7×10^−5^) standard deviations in inverse-normalized liver fat (Figure 1), whilst no difference was observed in non-Caucasian populations.

**Figure 1.**
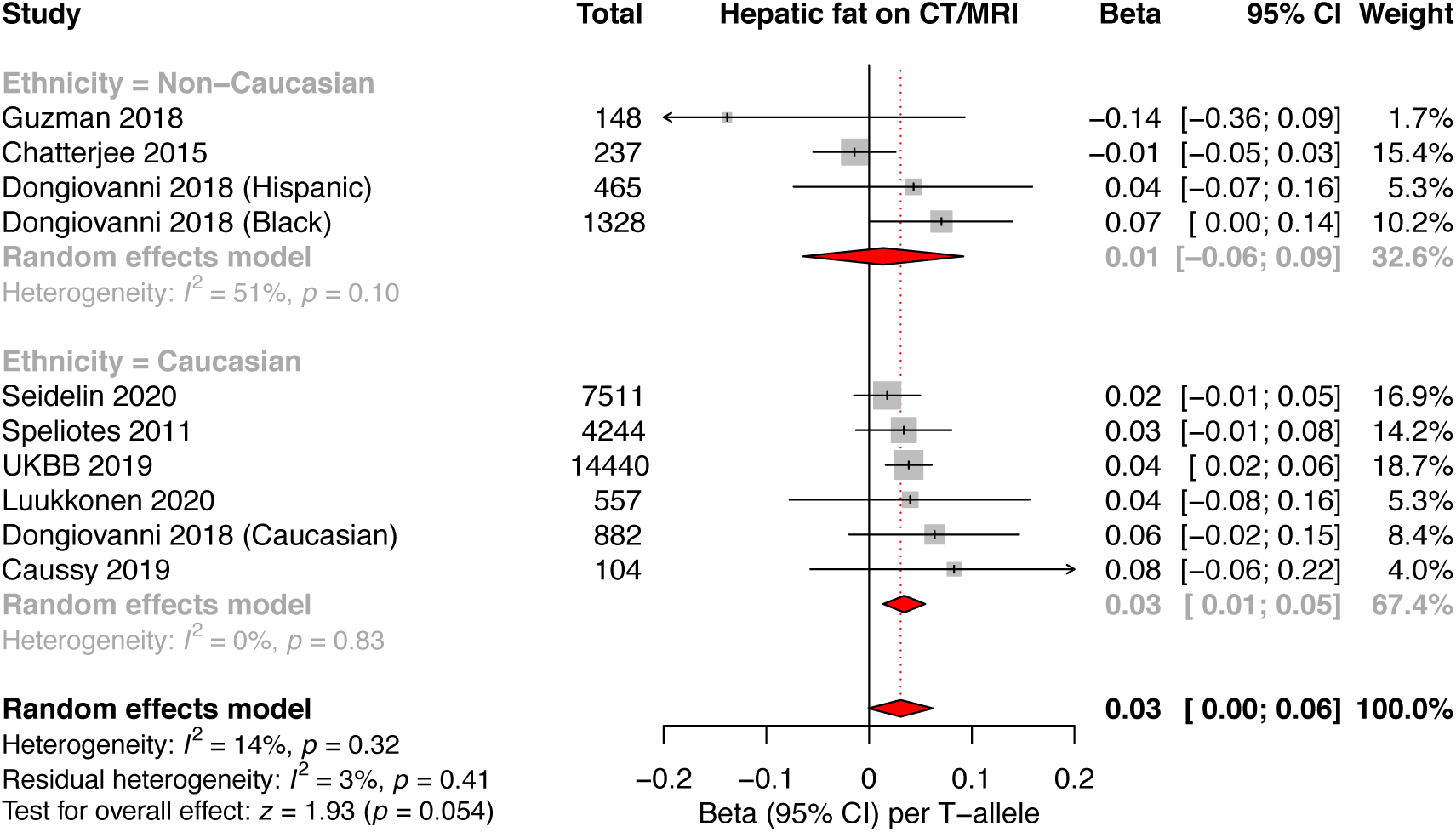
The effect of rs641738C>T on liver fat. Data from 29,916 individuals with CT or MRI liver fat. rs641738C>T positively associated with liver fat in Caucasian populations, where data represents standard deviation change in normalized liver fat per T-allele. CI, confidence interval; UKBB, UK BioBank.

A similar trend was observed using CAP and semi-quantitative ultrasound to assess steatosis severity in 12,224 adults (β 0.02 (95% CI -0.002, 0.04), Supplementary Figure 2).

rs641738C>T was also associated with NAFLD as a trait (OR 1.20 (95% CI 1.08, 1.32) using a recessive model of inheritance (Figure 2) but not using additive or dominant models (Supplementary Table 5). The effect was only observed in studies of Caucasian populations. The association remained after excluding three studies where there was a lack of similarity between cases and controls (OR 1.20 (95% CI 1.08, 1.33), I^2^ =18%, P_Q_=.26) using a recessive model of inheritance.

**Figure 2.**
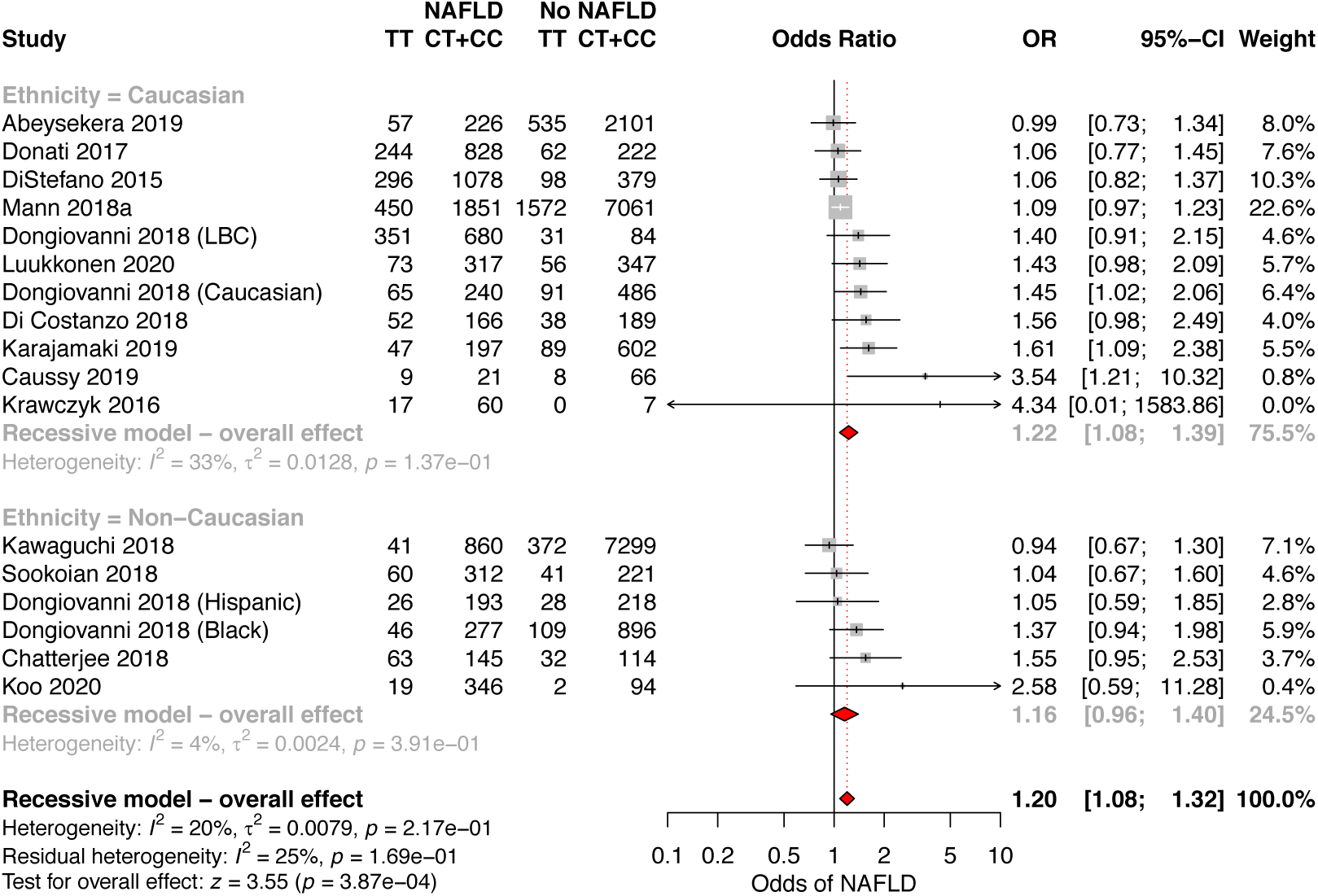
rs641738C>T is associated with higher odds of diagnosis of NAFLD. Data from 33,263 adults (9,713 cases and 23,550 controls) with radiologically defined steatosis for presence versus absence of NAFLD. CI, confidence interval; LBC, Liver Biopsy Cohort; OR, odds ratio.

However, Egger’s test suggested evidence of study distribution (publication) bias (p=0.01) and when using the Trim and Fill method to account for this bias, the positive association remained but was attenuated (OR 1.13 (95% CI 1.01, 1.26), P_z_=0.026, Supplementary Figure 3).

In patients with NAFLD, rs641738C>T was associated with the presence of severe steatosis (S1-2 vs. S3) on liver biopsy (OR 1.32 (95% CI 1.10, 1.59], Supplementary Figure 4), which was again due to a positive association in Caucasian individuals only (Table 2).

**Table 2.** Summary of results in adults from meta-analyses for dichotomous outcomes. Meta-analyses were performed using random effects with subgroup analysis for Caucasian and Non-Caucasian populations. Additive, recessive, and dominant genetic models were tested for all outcomes. Results using a recessive model of inheritance (CC+CT vs. TT) are shown for all outcomes, except for HCC, where a dominant model (CC vs. CT+TT) is shown. Due to use of three genetic models, critical p-value for effect summary is P_z_<0.017. Full results (with all genetic models) are in Supplementary Table 5. CI, confidence interval; HCC, hepatocellular carcinoma; NAFL, non-alcoholic fatty liver; NASH, non-alcoholic steatohepatitis; OR, odds ratio.

| Outcome | Genetic Model | Population | No. studies | Heterogeneity |  | Effect summary |  |  |  |
| --- | --- | --- | --- | --- | --- | --- | --- | --- | --- |
|  |  |  |  | I<sup>2</sup> (%) | PQ | OR | 95% CI |  | Pz |
| NAFLD diagnosis (control vs NAFLD) | Recessive | Overall | 17 | 20 | 0.27 | 1.20 | 1.08 | 1.32 | 4.0x10<sup>-4</sup> |
| NAFLD diagnosis (control vs NAFLD) | Recessive | Non-Caucasian | 6 | 4 | 0.39 | 1.16 | 0.96 | 1.40 | 0.12 |
| NAFLD diagnosis (control vs NAFLD) | Recessive | Caucasian | 11 | 33 | 0.19 | 1.22 | 1.08 | 1.39 | 0.0021 |
| Severe steatosis (S1-2 vs S3) | Recessive | Overall | 8 | 0 | 0.85 | 1.32 | 1.10 | 1.59 | 0.0032 |
| Severe steatosis (S1-2 vs S3) | Recessive | Non-Caucasian | 2 | 0 | 0.78 | 1.22 | 0.56 | 2.65 | 0.63 |
| Severe steatosis (S1-2 vs S3) | Recessive | Caucasian | 6 | 0 | 0.63 | 1.33 | 1.10 | 1.61 | 0.0034 |
| NASH (NAFL vs NASH) | Recessive | Overall | 9 | 0 | 0.60 | 1.20 | 1.04 | 1.39 | 0.013 |
| NASH (NAFL vs NASH) | Recessive | Non-Caucasian | 4 | 0 | 0.78 | 1.24 | 0.85 | 1.80 | 0.26 |
| NASH (NAFL vs NASH) | Recessive | Caucasian | 5 | 32 | 0.26 | 1.22 | 1.00 | 1.50 | 0.05 |
| Any fibrosis (F0 vs F1-4) | Recessive | Overall | 9 | 40 | 0.11 | 1.32 | 1.08 | 1.62 | 0.0073 |
| Any fibrosis (F0 vs F1-4) | Recessive | Non-Caucasian | 3 | 18 | 0.30 | 1.77 | 0.92 | 3.43 | 0.09 |
| Any fibrosis (F0 vs F1-4) | Recessive | Caucasian | 6 | 43 | 0.13 | 1.21 | 1.06 | 1.40 | 0.007 |
| Advanced fibrosis (F0-2 vs F3-4) | Recessive | Overall | 7 | 0 | 0.65 | 1.28 | 1.04 | 1.57 | 0.019 |
| Advanced fibrosis (F0-2 vs F3-4) | Recessive | Non-Caucasian | 2 | 0 | 0.64 | 0.96 | 0.50 | 1.85 | 0.90 |
| Advanced fibrosis (F0-2 vs F3-4) | Recessive | Caucasian | 5 | 0 | 0.52 | 1.32 | 1.06 | 1.63 | 0.011 |
| HCC (NAFLD vs NAFLD-HCC) | Dominant | Overall | 4 | 0 | 0.65 | 1.64 | 1.18 | 2.27 | 0.0031 |

### Histological NASH in adults

Data from nine studies (6,155 participants) found that rs641738C>T had a positive association with the presence of NASH on biopsy in adults (OR 1.20 (95% 1.04, 1.39), P_z_=0.013 Supplementary Figure 5).

### Fibrosis in adults

Liver biopsy data on presence of advanced fibrosis was available from seven studies (6,211 adults). Our primary outcome, presence of advanced fibrosis in adults (stage F0-2 versus stage F3-4), was positively associated with rs641738C>T in Caucasian populations (OR 1.32 (95% 1.06, 1.63), P_z_=0.011), though the effect was borderline when non-Caucasian populations were included (Figure 3). In addition, two studies used ICD-codes (International Statistical Classification of Diseases and Related Health Problems) in the UKBB cohort to identify individuals with NAFLD and advanced fibrosis or cirrhosis[51,83]. Both found positive associations below genome-wide significance: for example, using an additive model of inheritance Emdin *et al*. found the association between rs641738C>T and cirrhosis as β 1.15 (SE 0.13), using an additive genetic model.

**Figure 3.**
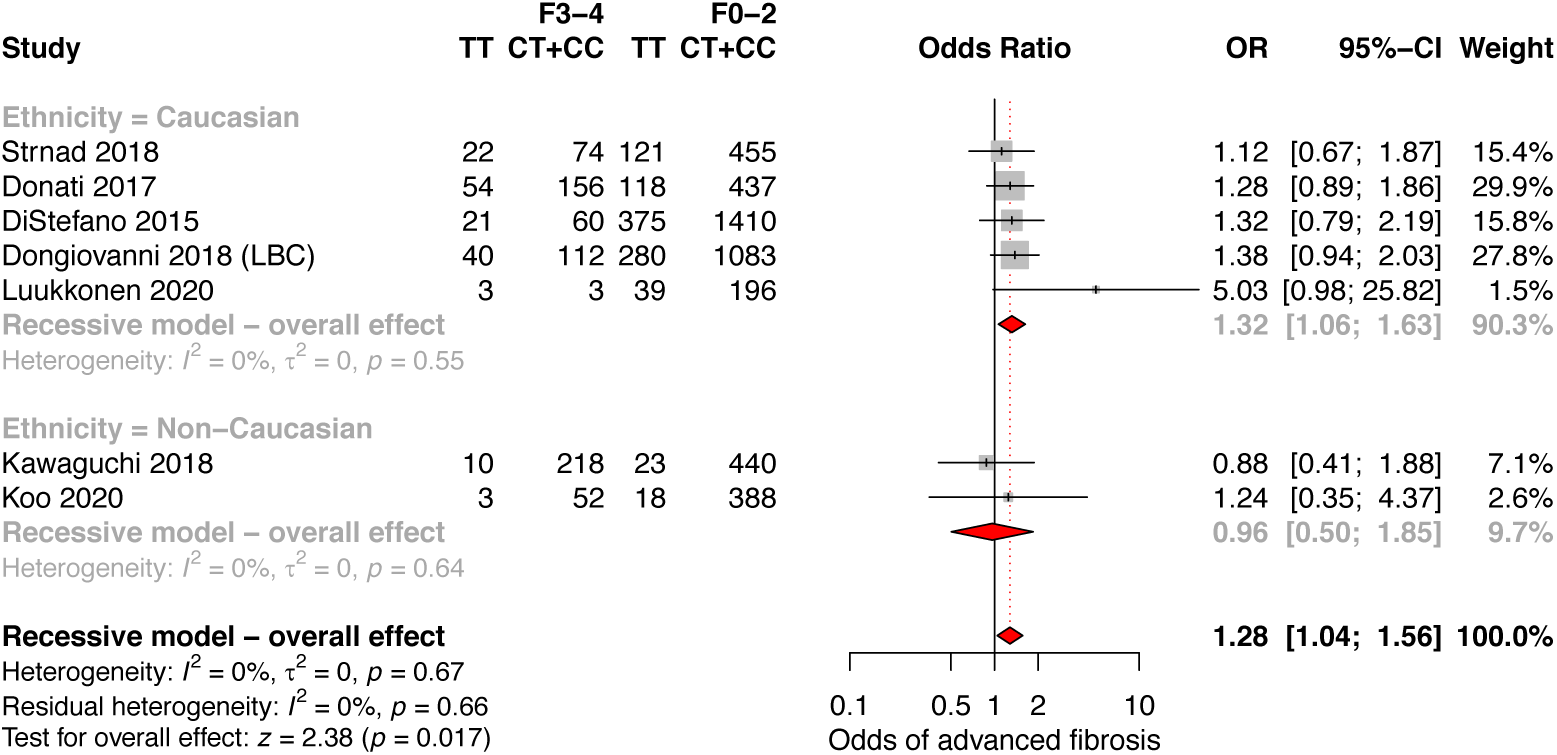
rs641738C>T is associated with higher odds of advanced fibrosis in Caucasian populations with NAFLD. Data from 6,211 adults (828 cases and 5,383 controls) with biopsy-proven NAFLD comparing advanced fibrosis (F3-4) versus F0-2, using a recessive model of inheritance (CC+CT vs. TT). CI, confidence interval; LBC, Liver Biopsy Cohort; OR, odds ratio.

Presence of any fibrosis (F0 versus F1-4) was also positively associated with rs641738C>T overall (OR 1.32 (95% 1.08, 1.62) and in Caucasian populations as a sub-group (Supplementary Figure 6).

### Development of hepatocellular carcinoma

Four cohorts (2,328 participants, 228 cases of NAFLD-HCC) reported on development of HCC in patients with NAFLD. rs641738C>T was associated with increased odds of HCC in NAFLD only when using a dominant model (CC vs. CT+TT) of inheritance (OR 1.64 (95% CI 1.18, 2.27), P_z_=0.003, Figure 4).

**Figure 4.**
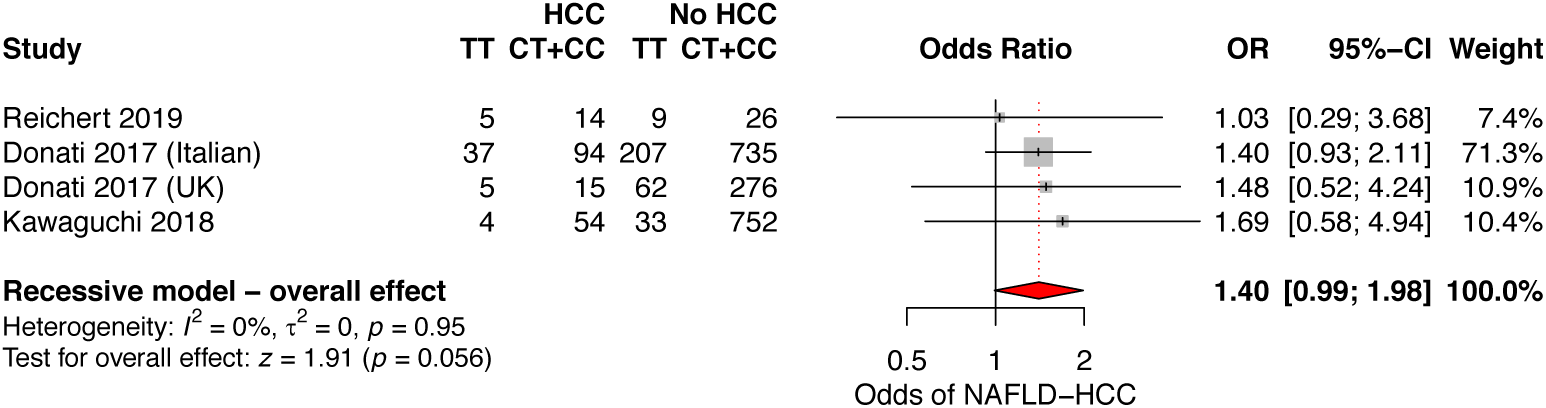
rs641738C>T is associated with higher odds of NAFLD-HCC in. Data from 2,328 adults with NAFLD assessing for the presence versus absence of HCC, using a dominant model of inheritance (CC vs. CT+TT).

### Effect on alanine aminotransferase (ALT)

Data from GWAS using log-transformed ALT (609,794 participants) were available for meta-analysis to investigate the role of rs641738C>T on ALT. The variant showed a positive association with ALT (β 0.004 (95% CI 0.002, 0.007), P_z_=0.002), which on sub-analysis was observed in Caucasian populations but not in non-Caucasian populations (Figure 5 & Supplementary Table 6).

**Figure 5.**
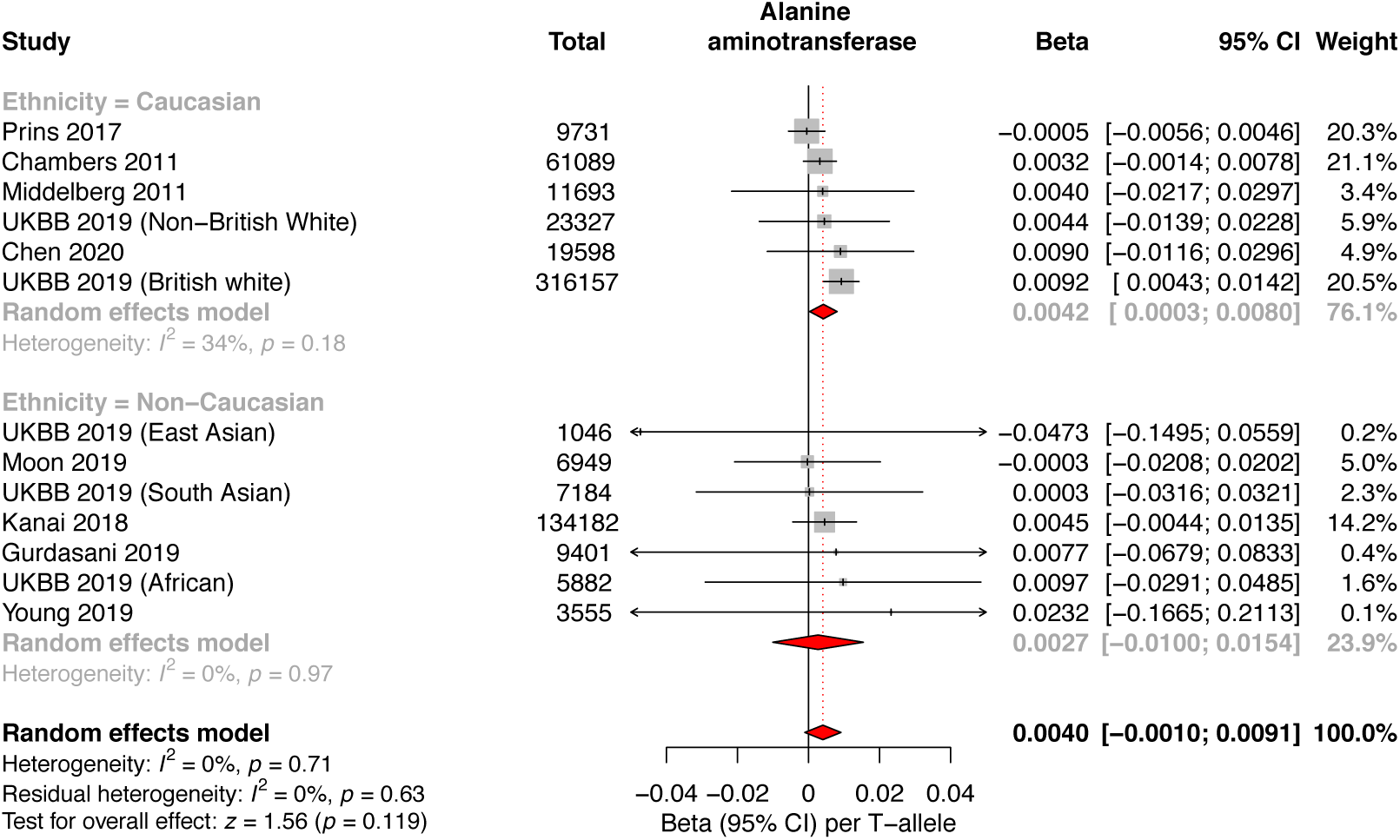
rs641738C>T is positively associated with alanine aminotransferase (ALT) in Caucasian populations in genome-wide association studies (GWAS). Meta-analysis of GWAS summary statistics from 609,794 participants for the association between rs641738C>T on logarithmically-transformed ALT using linear regression. CI, confidence interval; UKBB, UK BioBank.

Additionally, in the UKBB cohort, rs641738C>T was associated with a small, but statistically significant (P=2.0×10^−8^) increase in un-transformed ALT: 0.18 IU/L higher ALT per T-allele in this variant (Supplementary Table 7).

In the remaining cohort and case-control studies included in the meta-analysis (15,208 adults), rs641738C>T was not found to be associated with a change in ALT: mean difference between CC vs. TT 0.2 IU/L (95% CI -0.4, 0.8), Supplementary Table 8).

### Effect on serum lipids and insulin

Data from GWAS using log-transformed serum triglycerides (850,241 participants) found that rs641738C>T was associated with lower triglycerides (β -0.01 (95% CI -0.018, -0.006), P_z_=1.5×10^−4^), which on sub-analysis was observed in Caucasian populations but not in non-Caucasian populations (Supplementary Fig. 7). Similar findings were obtained from meta-analysis of mean difference in triglycerides between CC vs. TT -0.03 mmol/L (95% CI - 0.05, -0.01), Supplementary Table 8).

Data from GWAS (852,409 participants) found rs641738C>T to be positively associated with total cholesterol (β 0.007 (95% CI 0.003, 0.01), P_z_=2.3×10^−4^), in Caucasian populations but not in non-Caucasian populations (Supplementary Fig. 8). A borderline positive association was also observed in between rs641738C>T and high-density lipoprotein (HDL) cholesterol (β 0.009 (95% CI 0.001, 0.02), P_z_=0.02), Supplementary Table 6). There was no effect observed on low-density lipoprotein (LDL) or fasting insulin levels (Supplementary Tables 7 & 8).

### Effect of rs641738C>T on paediatric NAFLD

Data from ten studies (5,711 children) was used in the meta-analysis. rs641738C>T was not significantly associated with the diagnosis of NAFLD, liver fat content, or stage of liver histology (Supplementary table 9). The variant was observed to be associated with lower serum triglycerides in non-Caucasian children comparing CC vs. TT: -0.11 mmol/L (95% CI -0.14, -0.08).

## DISCUSSION

Identification of genetic variants associated with NAFLD has the potential to inform pre-clinical research and our understanding of hepatic metabolism. In this meta-analysis we have validated rs641738C>T near *MBOAT7* as a risk factor for the full spectrum of NAFLD in Caucasian adults.

A two-stage GWAS initially identified rs641738C>T as a genome-wide significant locus for alcohol-related cirrhosis[4]. *MBOAT7* was a potentially interesting target as an enzyme involved in (phospho)lipid metabolism, conceptually similar to other SNVs at GWAS-significance in alcoholic and non-alcoholic liver disease, namely *TM6SF2* and *PNPLA3*. Later studies found the variant to influence the full spectrum of fatty liver disease, from steatosis to NASH, to fibrosis, cirrhosis and HCC[5,18]. However, these associations have not been consistently replicated in the literature[20]. We conducted a meta-analysis to firmly establish the association of

### Main findings

We found that rs641738C>T was associated with higher liver fat content, higher ALT, and with higher odds of NAFLD diagnosis, severe steatosis, NASH, fibrosis, and HCC, particularly in Caucasian adults and in the homozygous ‘TT’ genotype. The effects sizes of rs641738C>T reported here are small compared to those of *PNPLA3* p.I148M and *TM6SF2* p.E167K, the two strongest steatogenic variants[3]. Also, the magnitude of change in alanine aminotransferase is small relative to that associated with variants in *PNPLA3, HSD17B13, MARC1*, and *TM6SF2*. The marginal positive effect on hepatic triglyceride content may suggest this variant acts through alterations in the composition of hepatic lipid, as well as quantity[18]. This is consistent with pre-clinical data on lipotoxicity, where the composition of hepatic fats influence development of NASH. On the other hand, a recent Mendelian randomization study using these variables as instruments to assess causality of fatty liver in determining fibrosis has shown the effect of steatosis highly correlates with fibrosis in all the genetic variables indicating that quantity of lipid rather than quality may be more important[56]. Functional studies are needed to understand the relationship between quality/quantity of fat and hepato-toxic/-protective mechanism in causing progression of disease.

The function of this variant is still relatively poorly understood and there is conflicting evidence as to whether rs641738C>T is associated with changes in hepatic expression of *MBOAT7*. Results from the GTEx Consortium show a strong negative association with T-allele[16], which is supported by data from Schadt *et al*.*[84]. MBOAT7* protein expression correlated with mRNA in liver biopsies from Mancina *et al*.*[5]* but this finding was not replicated by Sookoian *et al*.*[20]. MBOAT7* encodes LPIAT1, a 6 transmembrane domain protein involved in acyl-chain remodeling of membranes that influence intracellular membrane composition and circulating phosphatidylinositols[85]. Further, recent metabolite profiling data implicates *MBOAT7* as the causal gene for this SNV[66]. Moreover, TMC4 was found with a low expression in the liver[5] that is consistent with no mechanistic data supporting its role in NAFLD.

The hypothesis that *MBOAT7* is the causal gene underlying the association with liver disease at the locus is supported by the observation that mice deficient for *MBOAT7* have altered hepatic concentrations of polyunsaturated phosphatidylinositol[85]. Similarly, metabolite data from humans is strongly suggestive that rs641738C>T reduces MBOAT7 function[86]. In addition, two independent groups have found that loss of *MBOAT7* (but not *TMC4)* increases the severity of NAFLD in mice fed a high-fat diet[87,88].

These analyses suggest that rs641738C>T impacts the severity of NAFLD through a recessive model of inheritance, though some analyses (e.g. liver fat and ALT) were suggestive of a role using an additive genetic model. Other genetic variants are known to impact on all-cause mortality in a recessive manner, notably variants that perturb *HFE[83]*. Further mechanistic work is required to understand the extent to which the halo-insufficient state affects hepatocyte function.

We found no evidence of rs641738C>T on insulin resistance: the key driver of hepatic steatosis, as determined by unaltered fasting insulin concentrations. GWAS meta-analyses of type 2 diabetes have implicated p.I148M in *PNPLA3* and p.E167K in *TM6SF2* as significant risk loci (albeit with very modest effect size as compared to their effects on liver disease)[89] and Mendelian randomization studies indicate a causal role in determining insulin resistance mediated by the degree of liver damage[56,75]. Similarly, these two variants are associated with reduced risk of coronary artery disease; whilst our analysis did find lower serum triglycerides to be associated with this variant it has not been associated with lower rates of cardiovascular disease[90].

A strength of this meta-analysis is the large number of individuals with liver biopsy-derived phenotypic data as well as use of population-based GWAS data. The larger number of included studies and participants is likely to account for the different conclusions reached in this study compared to the previous meta-analysis by Xia *et al*.*[82]*.

### Limitations and quality of evidence

An important practical consideration is the population frequency of this variant in different ethnicities. The mean allelic frequency of the effect (T) allele is highly variable: from 0.24 in East Asians compared to 0.53 in those of South Asian ancestry[91]. Moreover, the majority of studies included in this meta-analysis used self-reported ethnicity, rather than genetic ancestry[92].

Though this analysis did include data from individuals of multiple ethnicities (and genetic ancestries) we only found evidence of an effect of this variant in Caucasian individuals. This is consistent with the initial discovery however, due to differences in patterns of linkage disequilibrium, we cannot exclude the possibility that a different nearby locus is associated with liver-related phenotypes in individuals of other genetic ancestries.

We found significant differences between adult and paediatric histological analyses. Whilst there were fewer clinical events (e.g. with advanced fibrosis) in children, the analyses did not show a trend congruous with those in adults. Paediatric NAFLD has a different histological phenotype to that of adults (with prominent periportal inflammation) and it is therefore plausible that this is a true lack of association in children with NAFLD.

The magnitude of effect observed across all associations is small in comparison to other well-established variants. The clinical relevance of rs738409C>G in *PNPLA3* has been validated with hard end-points[93] but large cohorts will be required to prospectively demonstrate the clinical risk associated with this variant near *MBOAT7*.

Though there was minimal heterogeneity across included studies, there was evidence of publication bias but the effect on diagnosis of NAFLD appeared to persist after attempting to account for this. Also of note, the numbers of individuals with NAFLD and HCC were comparatively low. It was also unique in only demonstrating an effect in the dominant, rather than recessive, model of inheritance. Further work in this area may improve the accuracy of effect estimates.

## Conclusions

rs641738C>T near *MBOAT7* is positively associated with liver fat, ALT, and histological severity in Caucasian adults with NAFLD, but negatively associated with serum triglycerides and with relatively small effect sizes throughout. These data validate this locus as significant in the pathogenesis of NAFLD.

## Data Availability

Summary statistics available upon request.

## ACKNOWLEDGEMENTS

The authors are grateful to the Raine Study participants and their families, and to the Raine Study research staff for cohort coordination and data collection. The authors gratefully acknowledge the following institutes for providing funding for Core Management of the Raine Study: The University of Western Australia (UWA), Curtin University, the Raine Medical Research Foundation, the UWA Faculty of Medicine, Dentistry and Health Sciences, the Telethon Kids Institute, the Women and Infants Research Foundation (King Edward Memorial Hospital) and Edith Cowan University).

We are extremely grateful to all the families who took part in the ALSPAC study, the midwives for their help in recruiting them, and the whole ALSPAC team, which includes interviewers, computer and laboratory technicians, clerical workers, research scientists, volunteers, managers, receptionists and nurses.

This study has been conducted using data from the Fenland study. The authors gratefully acknowledge the help of the MRC Epidemiology Unit Support Teams, including Field, Laboratory and Data Management Teams. The authors are grateful to the members of the EU-PNAFLD Registry, including Anita Vreugdenhil, Anna Alisi, Piotr Socha, Wojciech Jańczyk, Ulrich Baumann, Sanjay Rajwal, Indra van Mourik, Florence Lacaille, Myriam Dabbas, Deirdre A. Kelly, Quentin M. Anstee and the late Valerio Nobili. We would also like to thank Naga Chalasani for his helpful comments. This research has made use of the UK Biobank resource under project number 9914.

